# Knowledge, attitude, practice and fear of COVID-19: A cross-cultural study

**DOI:** 10.1101/2020.05.26.20113233

**Authors:** Mohammad Ali, Zakir Uddin, Palash Chandra Banik, Fatma A. Hegazy, Shamita Zaman, Abu Saleh Mohammed Ambia, Md. Kaoser Bin Siddique, Rezoana Islam, Fatema Khanam, Sayed Mohammad Bahalul, Md Ahiduzzaman Sharker, F M Akram Hossain, Gias U Ahsan

## Abstract

**Background:** Knowledge, Attitude and Practice (KAP), and Fear toward COVID-19 are an important issue when designing public health approaches to control the spread of this highly contagious disease like COVID-19 during the global pandemic period. Studies with KAP and fear measures are limited only regional or country level, not yet with global or cross-cultural populations. The study is aimed to measure KAP and fear level towards COVID-19 and explore its cross-cultural variances in knowledge by socio-demographic factors among the general population of 8 different countries over 5 continents.

**Method:** A cross-sectional online survey was conducted in April 2020 among 1296 participants using the Google form platform. Considering the social distancing formula and pandemic situation, we collect data using popular social media networks. Univariate and bivariate analyses were used to explore the collected data on KAP, fear, and sociodemographic factors.

**Result:** Overall knowledge score was 9.7 (out of 12) and gender differences (female vs male: 9.8 vs 9.5) were significant (p=0.008) in the bivariate analysis. Knowledge score variances found significant in some regions by gender, marital status and education qualification. The highest and lowest mean knowledge scores were recorded in the Middle East (10.0) and Europe (9.3). Despite having a high fear score (22.5 out of 35), 78.35% of respondents were in a positive attitude and 81.7% in good practice level. Fear score rankings: Middle East (1st; 23.8), Europe (2nd; 23.2); Africa (3rd; 22.7); South Asia (4th; 22.1); Oceania (5th; 21.9); and North America (6th; 21.7). We didn’t find a correlation between fear and knowledge.

**Limitation:** Due to the nature of the online survey, aged and rural populations are under-representing (e.g. more than half of the responders are 16-29 age group).

**Conclusion:** KAP and fear variation exist among geographical regions. Gender, marital status and education qualification are factors in knowledge variances for some regions. KAP and fear measures can assist health education programs considering some sociodemographic factors and regions during an outbreak of highly contagious disease and, which can uplift a positive attitude and good practice.

**Highlights:**

- Cross-cultural KAP and fear toward COVID-19 are evaluated
- Respondents from Europe scored less knowledge on COVID 19 but had more good knowledge level
- About 80% participants had positive attitude and good practice behavior.
- Interestingly, Participants in Oceania avoided more crowded places whereas, in Europe worn more masks during outing as a measure of prevention
- Participants in Middle East had the highest score in fear, and fear was independent of knowledge

## Introduction

COVID-19 disease caused by a novel coronavirus (SARS-CoV-2) has been declared as a pandemic by the World Health Organization (WHO) on March 11, 2020 (Bedford et al., 2020). Towards the end of April, it has been reported that the virus has spread across many countries around the world with fear-evoking death reports. (“Johns Hopkins Coronavirus Resource Center,” n.d.). This virus is highly contagious with some symptoms such as fever, dry cough, fatigue, myalgia, and dyspnea (B. L. Zhong et al., 2020). However, the human carrier of the virus without any clinical symptoms can also spread to other humans (Cascella et al., 2020). The mortality rate due to this virus is higher than common contagious diseases and can reach up to 15% (Baud et al., 2020). Due to these unique features, adequate management strategy is needed to battle the virus and safe life.

Effective pandemic management requires support from the population at risk and necessary measures to alleviate the spread of disease. Previous studies of a similar kind of contagious diseases, for example, the H1N1 flu outbreak, have revealed that Knowledge, Attitudes and Practices (KAP) play a significant role in personal experience and effect behaviors during pandemic (Yap et al., 2010). Other studies have shown that, improved knowledge has increased the uptake of preventive measures and has influence on the protective behavior at individual and community level (Lau et al., 2007; Leung et al., 2005).

KAP are the major adherence factors for the successful implementation of prevention and control measures for COVID-19 (Ajilore et al., 2017; Tachfouti et al., 2012). Knowledge and attitudes have greater impacts on emotions, personality, anxiety, panic and stress (Markel, 1999). These attributes also have influence on the preventive measures against infectious diseases and utilization of healthcare services due to fear and stigma (Person et al., 2004; Siddique et al., 2017). Studies on previous epidemics indicated that some people experience “fear” during a serious disease outbreak which requires support from public health professionals (Markel, 1999). Quarantine and isolation are necessary to reduce the spread of virus. There is evidence of increased fear during quarantine and isolation at the global and country level (Jefferson et al., 2008). In fact, “fear” is a psychophysiological construct linking to the neurobiology of consciousness (Adolphs, 2013). A person may fail to think rationally and clearly with the cognitive load when reacting to a pandemic with a higher level of fear. Individual’s fear of COVID-19 level may assist healthcare providers for further designing an appropriate program for better management.

Despite having a unique character of the COVID-19, the number of identified infected cases varied from country to country. Nonetheless, most of the countries around the world announced lock down as an emergency measures to ensure people stay at home and maintain social distancing. In spite of the same preventive measures taken by the governments of high-income countries, a number of countries in Europe and the USA have been seriously hit by COVID-19 pandemic. However, low and middle-income countries such as Bangladesh and Egypt reported fewer identified cases (“Johns Hopkins Coronavirus Resource Center,” n.d.). Therefore, it would be interesting to consider whether KAP and fear levels are different between high-income and middle-income countries. There is a research gap combining KAP and fear towards COVID-19. The novelty of this study is exploring cross-cultural KAP and fear issues by collecting data from 8 different countries over 5 continents around the globe during the peak global pandemic period in April. This study aimed to (1) measure KAP and fear level towards COVID-19 and explore its cross-cultural variances among the general population of Australia, Bangladesh, Canada, Egypt, Sweden, UAE, UK and the USA, (2) examine the effect of cross-cultural knowledge level towards COVID-19 on socio-demographic factors (e.g. age, gender, marital status, educational qualification and occupational status).

## Methods and materials

### Study settings and participants

An online cross-sectional survey was conducted from April 6-27, 2020, using a ‘Google Form’ platform. Social media platforms such as Facebook, Facebook Messenger, WhatsApp and Skype were used to data collection because of COVID-19 pandemic situation. These platforms are popular around the world and a convenient platform to reach a large number of people within a short time. The survey invitation was sent to the social media contacts with the Authors living in different countries of the world. This Google form was an open invitation to social media users, and the posts were repeated every three days as a reminder to the participants. We could not calculate the response rate as it was simultaneously shared by the authors and their social media friends. Our participants include both male and female population aged 16 years and above and willingly took part in the survey. Current or recovered COVID-19 patients, health-care workers/volunteers who are working in management or control program were excluded. Respondents from other than eight chosen countries were also excluded. The survey form contained a brief introduction to the background, objective, and procedures. The survey was voluntary, anonymous and confidential. We got the response from 1296 invitees and after considering inclusion and exclusion criteria, we include 1255 participants for data analysis.

The ethical approval was taken from the Ethics Committee of North South University, Bangladesh (the institutional review board registration number: NSU-IRB-20-013023). Informed consent was taken using the ‘Google Form’ platform before data collection. All the concerns related to biomedical research according to the Helsinki declaration were followed strictly throughout the study.

### Measurements

The online survey questionnaire had three parts. The *first part* consisted of socio-demographic variables, including age, gender, marital status, education, occupation, and country of current residence.

In the *second part*, we used a questionnaire to assess the knowledge, attitude, and practice toward COVID-19 developed by Bao-Liang Zhong et. al. (B. L. Zhong et al., 2020). This questionnaire had 16 individual questions; among them, the first 12 questions represented the knowledge part. These 12 questions were answered on a true/false basis with an additional option of “I don’t know”. A correct answer was given 1 point and an incorrect/unknown answer was given 0 points. The total knowledge score ranged from 0 to 12, with a higher score explaining a better knowledge of COVID-19. The middle two questions were used to measure attitude toward COVID-19 which was scored the same as knowledge questions. The last two questions with yes/no answers were used to assess the respondents’ practice toward COVID-19. The level of knowledge was measured using the Bell curved approach (mean±1SD). The level of knowledge was classified as ‘Poor’ (less than Mean-1SD); ‘Average’ (Mean-SD to Mean+1SD); and ‘Good’ (Mean+1SD).

In the *third part*, fear was measured by the Fear of Coronavirus-19 Scale (FCV-19S) developed by Ahorsu et al. (Ahorsu et al., 2020). 7 items were included in the FCV-19S (e.g. “I am most afraid of coronavirus-19”) measuring the fear of the respondents on COVID-19. Participants were asked to rate their agreement with the statement using a 5-point Likert’ scale and the following options and the scores were 1= ‘Strongly Disagree’, 2= ‘Disagree’, 3= ‘Neither Agree and/or Nor Disagree’ 4= ‘Agree’, and ‘5 = Strongly Agree’. The minimum score for each question was 1, and the maximum was 5. A total score was calculated by adding up each item score (ranging from 7 to 35). The higher the score, considered the greater the fear of coronavirus-19. The scale has a robust psychometric property. It’s a reliable (α = .82 and ICC = .72) and valid scale for general population in assessing fear of COVID-19 (Ahorsu et al., 2020).

### Statistical Analysis

First of all, data was managed and cleaned as well as logically checked for internal consistency. Some of the data were discarded for their incompleteness or their inconsistency. The statistical analysis part was divided into two fundamental parts. The first part was the *univariate analysis* where the frequency, percentage, mean, and standard deviation (SD) were done as appropriate to describe the knowledge, attitudes, practice, and fear scores according to the sociodemographic characteristics of the respondents. The second part was the *bivariate analysis* where the independent sample’s t-test, one-way analysis of variance (one-way ANOVA), were done as appropriate to see the mean difference within the groups. Data analyses were conducted with SPSS software version 25.0. The statistical significance level was set at p < 0.05 (two-tailed). For data analysis, we assigned the respondents according to their greater continental/sub continental geographical region: South Asia (Bangladesh), Oceania (Australia) North America (Canada and USA), Europe (Sweden and UK), Middle East (UAE) Africa (Egypt). To explore the cross-cultural issue, all the data is presented into the tables and graphs by the area of residence of the respondents.

## Results

### Trajectories of “Knowledge” score by Cross-cultural Socio-demographic factors

Table 1 shows the sociodemographic characteristics in comparison to the knowledge score. Overall male and female ratios were 1:1.1 and more than half of the respondents were in the age group of 16 to 29 years (52.6%) and married (51.2%). Majority of (84.6%) of the respondents’ educational qualification was graduation and above. Almost three out of five respondents were employed (58.8%) in any way. The total mean knowledge score was 9.6 (out of 12) whereas, the highest and lowest mean knowledge score were recorded in Middle East (10.0) and Europe (9.3) respectively.

We found some variances in knowledge by sociodemographic factors. The overall mean knowledge score by (1) *age* was 9.6 whereas the highest and lowest mean knowledge score were recorded in Middle East (10.2) and South Asia (9.2) respectively, but the difference was not significant within the age group of the inter region; (2) *gender* was 9.7 whereas the highest and lowest mean knowledge scores were recorded in the Middle East (9.9) and European (9.3) region respectively and the male and female knowledge score difference was significant for overall (p = 0.008) and Oceania (p = 0.007) region; (3) *marital status* was 9.6 whereas the highest and lowest mean knowledge scores were recorded in Middle East (10.0) and Europe (9.1) respectively and the knowledge scores difference among the married, never married and others was significant for North American (p=0.007); (4) *educational qualification* was 9.3 and it was significant (p < 0.001) whereas the highest and lowest mean knowledge score was recorded in Middle East (9.7) and Europe (8.9) respectively and the knowledge score difference between different level of education was significant for South Asia (p < 0.001), Oceania (p = 0.025), Meddle East (p = 0.009) and African (p = 0.036) regions; (5) *occupational status* was 9.6 whereas the highest and lowest mean knowledge score was recorded in Europe (9.9) and Middle East (9.0) respectively and the difference was not significant for their different types of jobs.

**Table 1.**
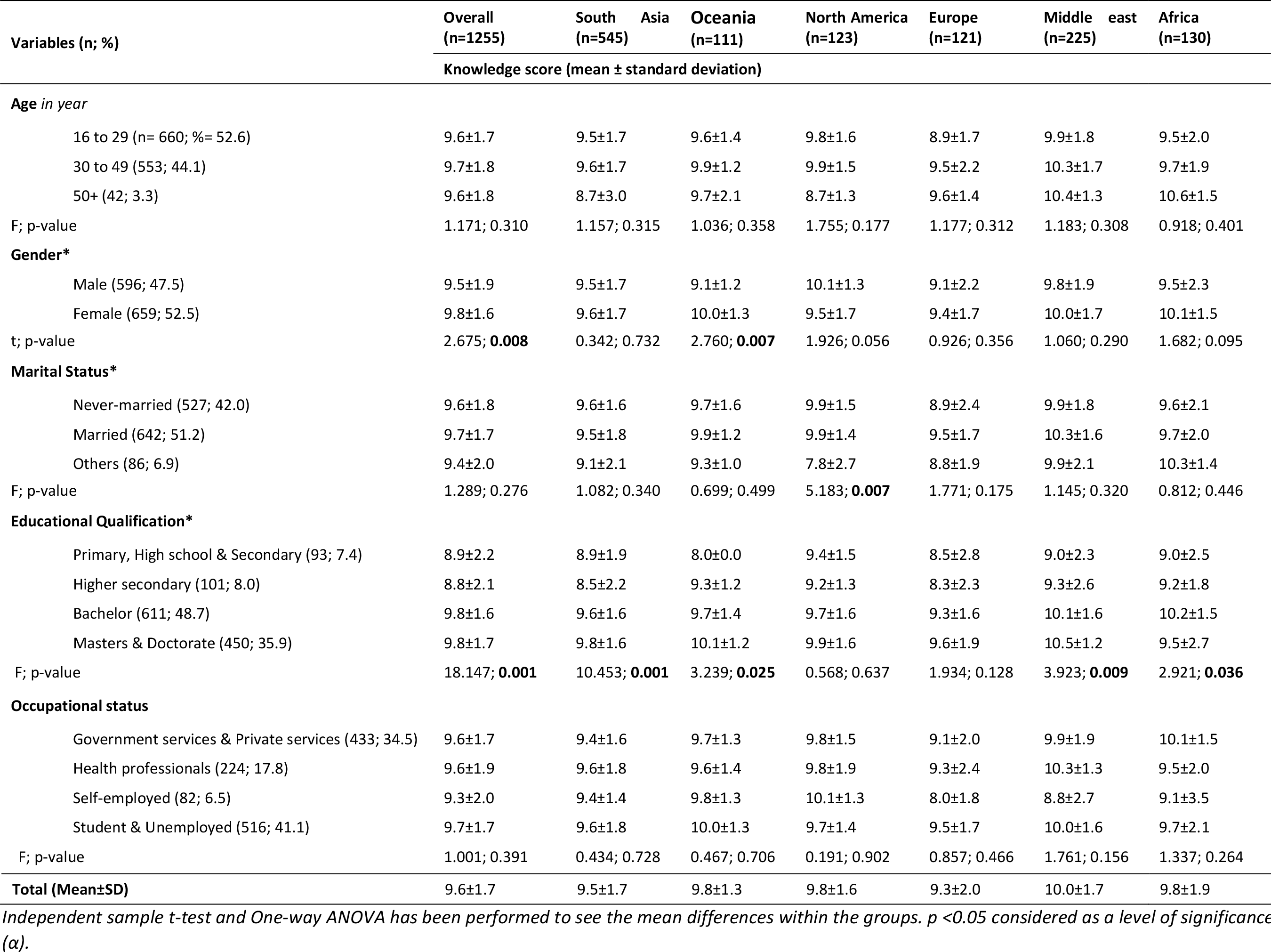
Overall characteristics of the respondents by their knowledge score and area of residence.

### Percentage of correct responses and level of “Knowledge” by area of residence

Figure 1 shows the distribution of correct responses by geographical area. The mean (±SD) of the total score was 9.7 (±1.7) out of 12 possible knowledge score. We classified knowledge score as poor (less than Mean-SD), average (Mean-SD to Mean+SD) and good (more than Mean+SD). Therefore, for “poor knowledge” the cut off was less than 8.0; for “average knowledge” the cut off was 8.0-11.4; and for the “good knowledge” the cut off was more than 11.4. The highest percentage of respondents with good knowledge was reported in Europe (18.2%) region whereas lowest was reported in Oceania (2.7%) and the difference was 15.5%; In addition, highest percentage of respondents with average knowledge was reported in Oceania (86.5%) region whereas lowest was reported in Africa (75.7%) and the difference was 10.8%; as well as, highest percentage of respondents with poor knowledge was reported in Africa (14.2%) region whereas lowest was reported in Europe (5.0%) and the difference was 9.2%.

**Figure 1:**
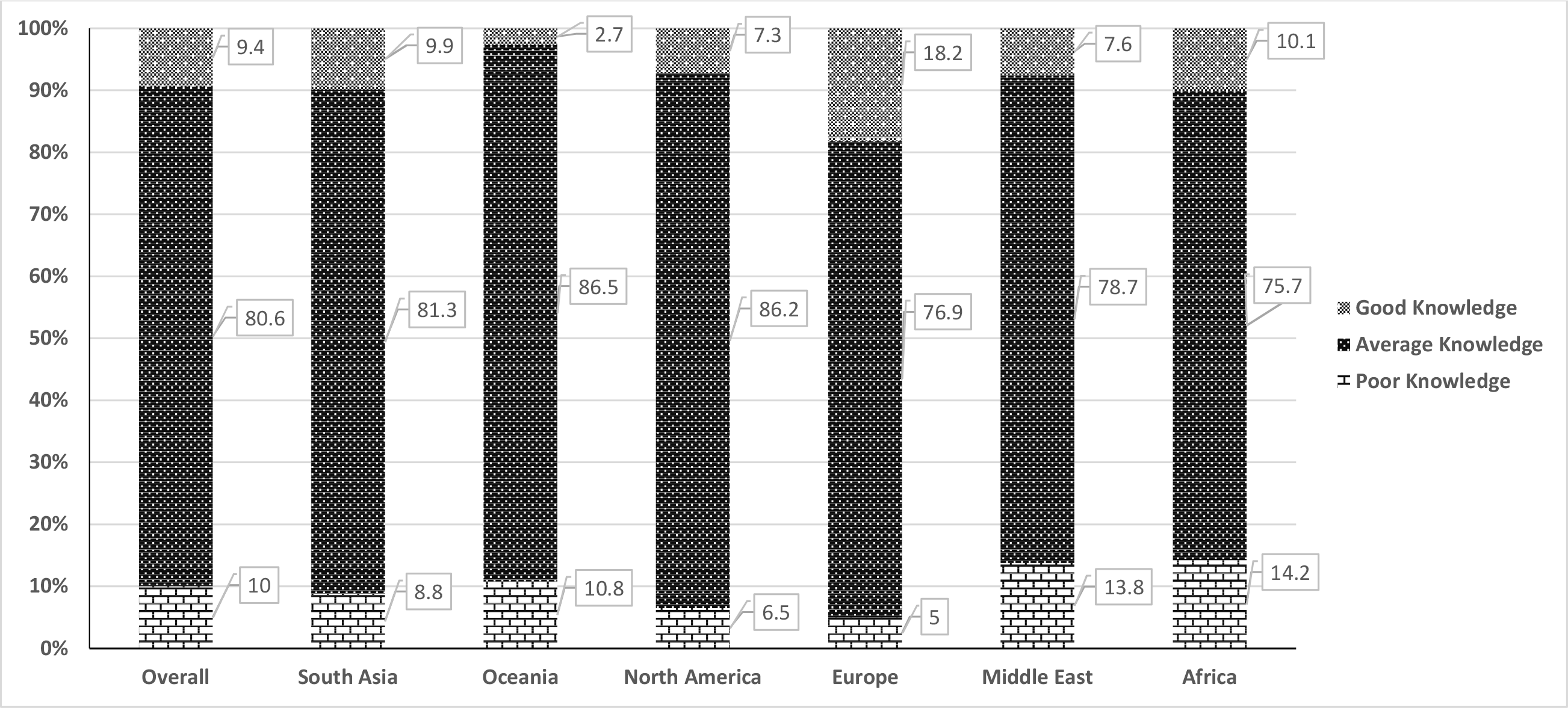
Level of knowledge according classified knowledge score by geographical area (n=1255)

### Percentage of correct responses of “Attitudes” and “Practices” by area of residence

The highest correct response for the 1^st^ question regarding attitudes was recorded in North America (93.5%) and the lowest correct response was recorded in Africa (83.8%) region and the difference was 9.7%. The highest correct response for the 2^nd^ question regarding attitudes was recorded in North America (75.6%) and lowest correct response was recorded in South Asia (62.0%) region and the difference were 13.6% (Figure 2).

**Figure 2:**
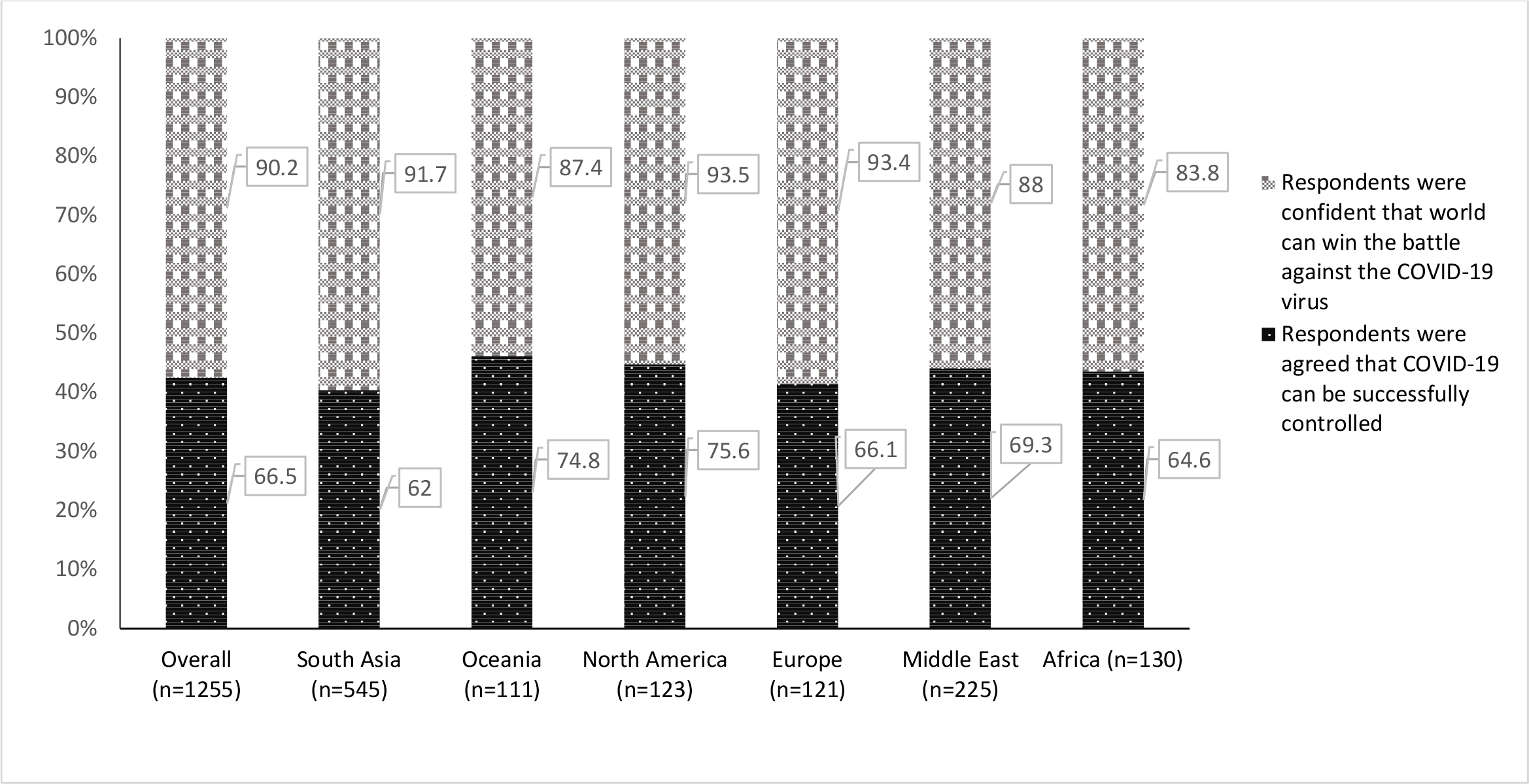
Attitudes of the respondents towards COVID 19 according to the area of residence.

The highest correct response was recorded in South Asia (97.6%) and lowest correct response was recorded in Oceania (47.7%) region and the difference was significant (49.9%). The second question was, whether the respondents avoided the crowded places? And the highest correct response was recorded in the Oceania (93.7%) and lowest correct response was recorded in the Africa (74.6%) region and the difference was 19.1% (Figure 3).

**Figure 3:**
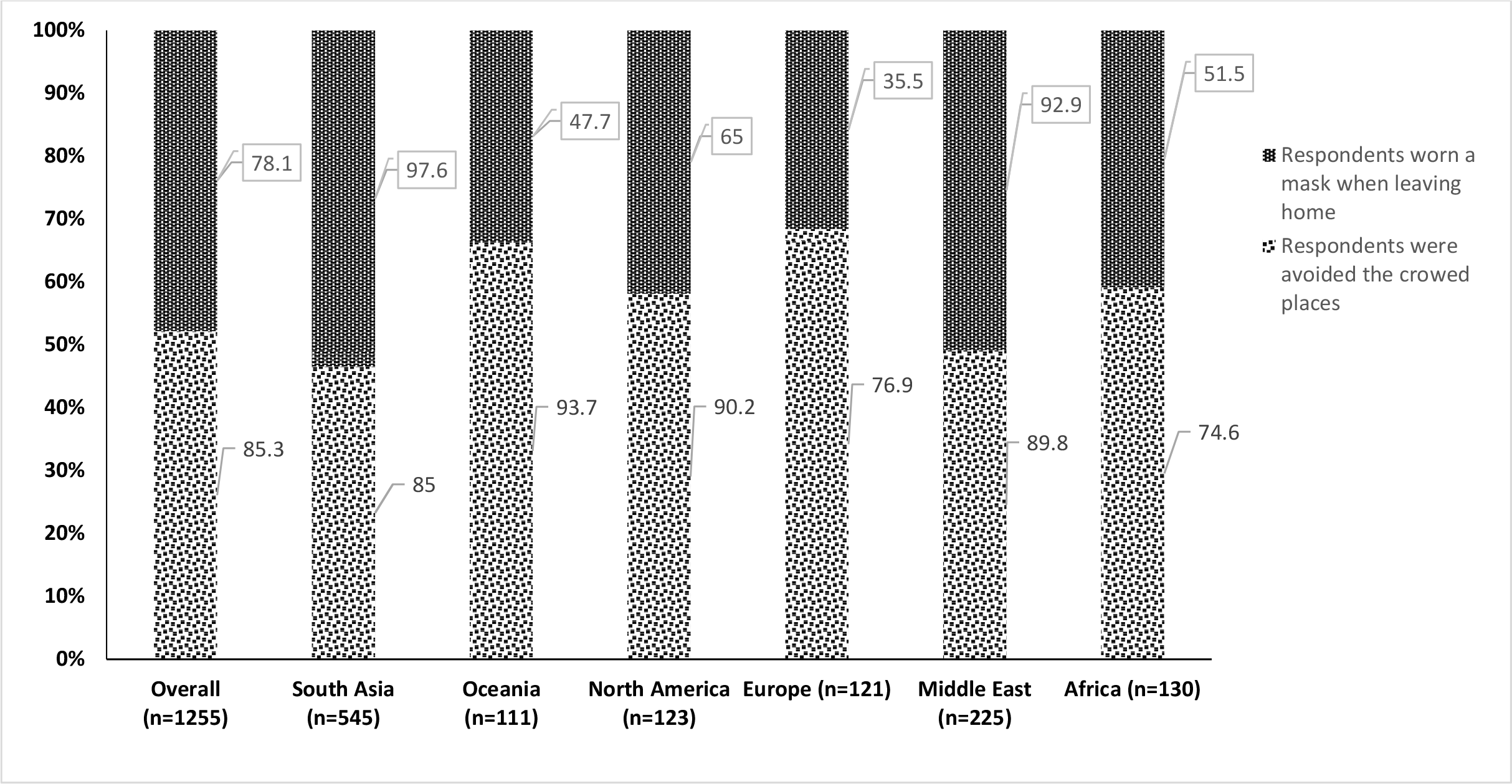
Practices of the respondents towards COVID 19 according to the area of residence.

### “Fear” by geographical region

Regarding the fear related to COVID-19, three out of ten (30.5%) respondents were positively responses about fear. Respondents from South Asia reported highest positive responses in fear (34.8%) whereas, respondents from Middle East reported lowest positive responses in fear (22.7%) and the response difference was 12.1%. When we assessed their mean score for the fear, we found that the highest mean score for fear was in Middle East (Ranked, 1st; 23.8±5.5) which indicates greater fear of cororonavirus-19 compared to other’s continents like Europe (Ranked, 2nd; 23.2±5.8); Africa (Ranked, 3rd; 22.7±5.0); South Asia (Ranked, 4th; 22.1±5.7); Oceania (Ranked, 5th; 21.9±5.8); and North America (Ranked, 6th; 21.7±5.5), but the difference was minimum (Table 2).

**Table 2:** Fear regarding COVID-19 by country of residence among the respondents.

| Variables | Overall (n=1255) | South Asia (n=545) | Oceania (n=111) | North America (n=123) | Europe (n=121) | Middle East (n=225) | Africa (n=130) |
| --- | --- | --- | --- | --- | --- | --- | --- |
| **\*Distribution of respondents by positive responses on fear about COVID 19; % (95% Confidence Interval)** | 30.5 (29.7 to 36.9) | 34.8 (30.8 to 38.8) | 34.6 (25.8 to 43.4) | 33.8 (25.4 to 42.2) | 32.7 (24.3 to 41.1) | 22.7 (17.2 to 28.2) | 24.1 (16.7 to 31.5) |
| **\*\*Fear of Coronavirus-19 Scale** | *(Mean ±SD) out of total score 5* |  |  |  |  |  |  |
| I am most afraid of coronavirus-19 | 2.6±1.1 | 2.4±1.0 | 2.5±1.1 | 2.6±1.2 | 2.5±1.1 | 2.8±1.0 | 2.9±1.0 |
| It makes me uncomfortable to think about coronavirus-19 | 2.7±1.1 | 2.7±1.2 | 2.6±1.1 | 2.7±1.0 | 2.7±1.1 | 2.8±1.0 | 2.6±1.0 |
| My hands become clammy when I think about coronavirus-19. | 3.8±1.0 | 3.8±1.0 | 3.7±0.9 | 3.6±1.0 | 3.9±1.0 | 3.8±1.0 | 3.5±1.0 |
| I am afraid of losing my life because of coronavirus-19 | 3.3±1.1 | 3.2±1.1 | 3.2±1.2 | 3.1±1.0 | 3.4±1.2 | 3.4±1.2 | 3.3±1.0 |
| When watching news and stories about coronavirus-19 on social media, I become nervous or anxious | 2.8±1.1 | 2.8±1.1 | 2.5±1.0 | 2.6±1.0 | 2.9±1.1 | 3.0±1.1 | 2.8±1.0 |
| I cannot sleep because I'm worrying about getting coronavirus-19 | 3.9±1.0 | 3.8±1.1 | 4.0±1.0 | 3.7±1.1 | 4.0±1.1 | 4.1±0.9 | 4.1±0.9 |
| My heart races or palpitates when I think about getting coronavirus-19 | 3.6±1.1 | 3.5±1.1 | 3.5±1.2 | 3.5±1.1 | 3.8±1.1 | 3.9±1.1 | 3.7±1.1 |
| **Total (Mean±SD; Minimum to Maximum) score out of total score 35** | **22.5±5.6; 7 to 35** | 22.1±5.7; 7 to 35 | 21.9±5.8; 7 to 34 | 21.7±5.5; 7 to 35 | 23.2±5.8; 9 to 35 | 23.8±5.5; 7 to 35 | 22.7±5.0; 9 to 35 |
| **Fear Ranking position** |  | 4<sup>th</sup> | 5<sup>th</sup> | 6<sup>th</sup> | 2<sup>nd</sup> | 1<sup>st</sup> | 3<sup>rd</sup> |

\*"Agreed" or "Strongly agreed" responses considered as positive responses for fear
\*\*The participants level of agreement based on fear scale. A total score was calculated by adding up each item score (7 x 5 =35 maximum possible).

### Relationship between “Knowledge” and “Fear”

Correlation patterns between knowledge score and the fear score are demonstrated in figure 4. We didn’t see any correlation between knowledge and fear in this study data.

**Figure 4:**
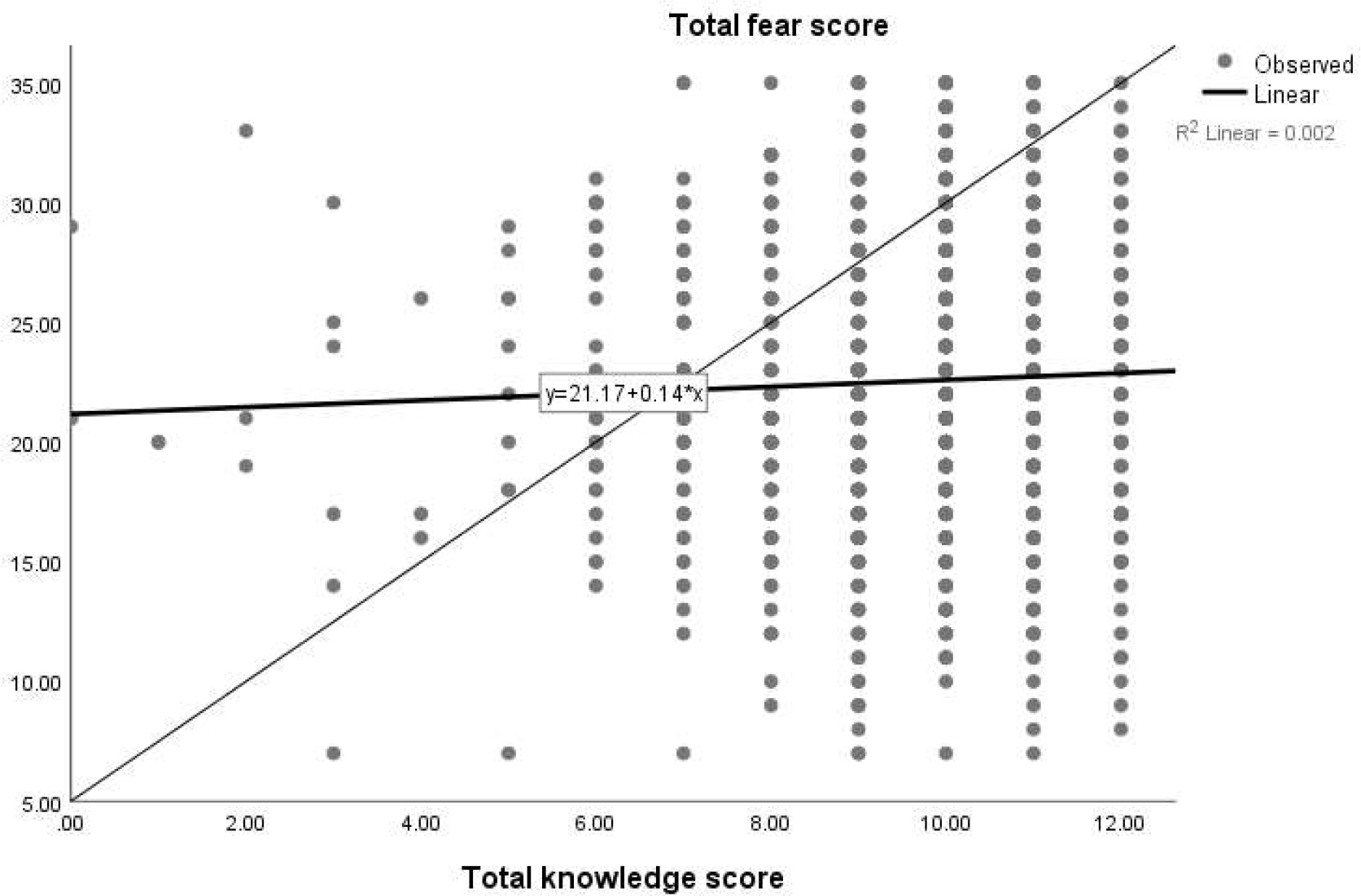
Correlation between knowledge score and the fear score of the respondents (n=1255)

## Discussion

In our study, a mixed knowledge score was observed for different demographic variables within the different regions. However, the knowledge score mean was relatively better in Middle East and relatively poor in European region. The marital status, occupation and gender factors were associated to knowledge in the bivariate analysis. South Asian respondents had highest positive responses in fear, but they ranked 4^th^. Interestingly, respondents from Middle East reported lowest positive responses in fear, but they ranked 1^st^ by the Fear level. This indicates that the respondents from Middle East had the greater fear on COVID-19. We didn’t find a correlation between knowledge and fear score. This signifies that fear is not dependent on the respondent’s knowledge on COVID-19.

In this study, the majority of the participants were young adults and middle-aged and more than half of the respondents were female. Interestingly, knowledge score towards COVID-19 was higher among the female and overall score were statistically significant. In Oceania region, level of knowledge of females has been found to be better than males followed by Africa, the Middle East, Europe and South Asia. However, in North America, the level of knowledge has been found to be higher than females. Similar to our study, Chinese’s females scored higher in the knowledge section when a KAP study towards COVID-19 conducted in China (B.-L. Zhong et al., 2020). Interestingly, according to the recent statistics, there is higher prevalence of male patients than females (Jin et al., 2020).

Our study was dominated by the educated population. Predictably, knowledge regarding COVID-19 was higher in the graduate and postgraduate participants. Especially in the South Asian and Oceania region, participants who did not attend university are scored lower. These findings were consistent with other previous studies where the authors found poor knowledge about the infectious disease among non-graduate population (Wang et al., 2018). We found significant differences in overall and Middle east regional knowledge scores by educational status.

Previous studies found that the spread rate of infectious diseases is higher among the population who did not have good knowledge about the disease (Goni et al., 2019; Rassi et al., 2019). In our study we found lowest knowledge score among participants of highly affected Europe region. The comparative picture of the rate of COVID-19 infection during our survey period (“Johns Hopkins Coronavirus Resource Center,” n.d.) and knowledge level distribution in our result (e.g. good, medium and poor) is somehow mixed. The reason might be the nature of the previous infectious disease conditions and this global pandemic condition is quite different. Participant’s regional social norm or cultural issue on mass media exposure might be so different for getting information on knowledge about this pandemic.

In China, the majority of the respondents held an optimistic attitude towards the COVID-19 epidemic: 90.8% believed that COVID-19 will be successfully controlled, and 97.1% had confidence that China can win the battle against the virus (B.-L. Zhong et al., 2020). Nonetheless, when we measure the attitude towards this pandemic, we found, 90.2% of the respondents were confident that the world will win the battle against COVID-19, however, overall only 60% of participants were optimistic that this disease can be successfully controlled. The region-wise optimistic attitude was almost the same.

Despite a severe attack by COVID-19 in Europe, we found only 35.5% of Europeans wore a mask when going outside. This rate was significantly lower than the overall practice rate (78.1%) of wearing masks when they went outside. Wearing mask rate was also considerably lower among participants living in another severely affected region of North America. As a matter of fact, our study result supports the reality of limiting contagious disease by good practice behaviors (Squibb and Yardley, 1999).

Previous scientific study has shown that fear of disease negatively affected public health efforts to control the disease outbreak. (Person et al., 2004). In our study, we found a higher fear score among participants from the Middle East and followed by Europe and Africa. The highest positive responses in fear found in South Asia. Region-specific cultural issue and mass media exposure frequency (e.g. news on death due to COVIT 19) might be a reason for this fear. However, we didn’t find a correlation between knowledge and fear score of the respondents in our study.

*Limitations* of the online survey cannot be ruled out in our study. Due to limited access to the internet, old age and rural people who have been more likely to have poor KAP were not included in the study. Selection bias may be significant if those of poor level KAP people appear to have less participation. In fact, KAP for vulnerable populations deserves special research attention. Moreover, there is a risk of bias due to online data collection. Ethical challenge is a fact in the cross-cultural study (Durham, 2014). However, we have mitigated the challenge for this study by maintaining some measures, such as (1) collecting informed consent before participating in the survey, (2) depersonalizing responders and (3) ensuring confidentiality for better personal security measure like other surveys in public health (Survey to Identify the Public Health Ethics Needs of Public Health Practitioners in Canada: Preliminary Results, n.d.).

Despite having those limitations, the cross-cultural nature is the main strength of this study. It’s difficult to compare this study to other similar kinds of studies. To our knowledge, no study measured the cross-cultural value of KAP and fear of COVID-19. Therefore, the result of this study is a novel contribution in the scientific literature. This study imposes a chance to evaluate the relationship between KAP and infection numbers among the population in a different continent. A representative from several professions and all ages made the study more generalized.

The majority of the participants in this study shows “average” level of knowledge (overall 80.6% according to the knowledge level classification of this study). Future cross-cultural study is needed to clarify the fact and find a way how to improve a “good” level of knowledge. It will be helpful to guide better educational health campaign strategies targeting global people by considering cultural/regional aspects. This study suggesting that mass media health education programs aimed at improving knowledge and reducing fear could help encourage an optimistic attitude and maintain safe practices that might result in controlled infection rate by creating awareness among global population considering their culture or regional social norm. KAP and fear measures can assist health education programs considering some sociodemographic factors and geographical regions during an outbreak of highly contagious disease.

## Data Availability

The data sets used and analyzed for this study are available. Please feel free to ask to the corresponding author. We have also provided some supplementary data.

## Declarations

### Ethical approval

We conducted the study according to the guidelines laid down in the Declaration of Helsinki and all procedures involving human subjects were approved by the Institutional Review Board (IRB) of North South University (NSU-IRB-20-013023). Informed consent was taken using the ‘Google Form’ platform before participating the online survey. The survey was voluntary, anonymous and confidential.

### Consent to Publish

Not applicable.

### Author’s contributions

Mohammad Ali: Conceptualization, Methodology, Investigation, Data curation, Data analysis conceptualization, Writing - original draft, Writing - review & editing, Validation. Zakir Uddin: Conceptualization, Investigation, Formal data analysis, Writing - review & editing, Supervision, Validation. Palash Chandra Banik: Data curation, Formal data analysis, Writing-result interpretation draft, Writing-review & editing, Validation. Fatma A. Hegazy: Writing - review, Validation. Shamita Zaman: Writing - English language usage, grammar, and spelling, review and editing, Validation. Abu Saleh Mohammed Ambia: Writing - English language usage, grammar, and spelling, review and editing, Validation. Md. Kaoser Bin Siddique: Writing-introduction draft, Writing - review, Validation. Rezoana Islam: Writing - review, Validation. Fatema Khanam: Writing-review, Validation. Sayed Mohammad Bahalul: Writing-review, Validation. Md Ahiduzzaman Sharker: Writing-review, Validation. F M Akram Hossain: Writing-review, Validation. Gias U Ahsan: Supervision, Validation.

## Acknowledgements

All the authors acknowledge the participants for providing us the information to conduct the study. Authors also would like to thank **Hasan Hafizur Rahman** from Canada, **Ifty Mahmud** from Australia and **Md. Ahsan Ullah Shah Masud** from Bangladesh for helping in data collection.

## SUPPLIMENARY DATA

**Table 4.**
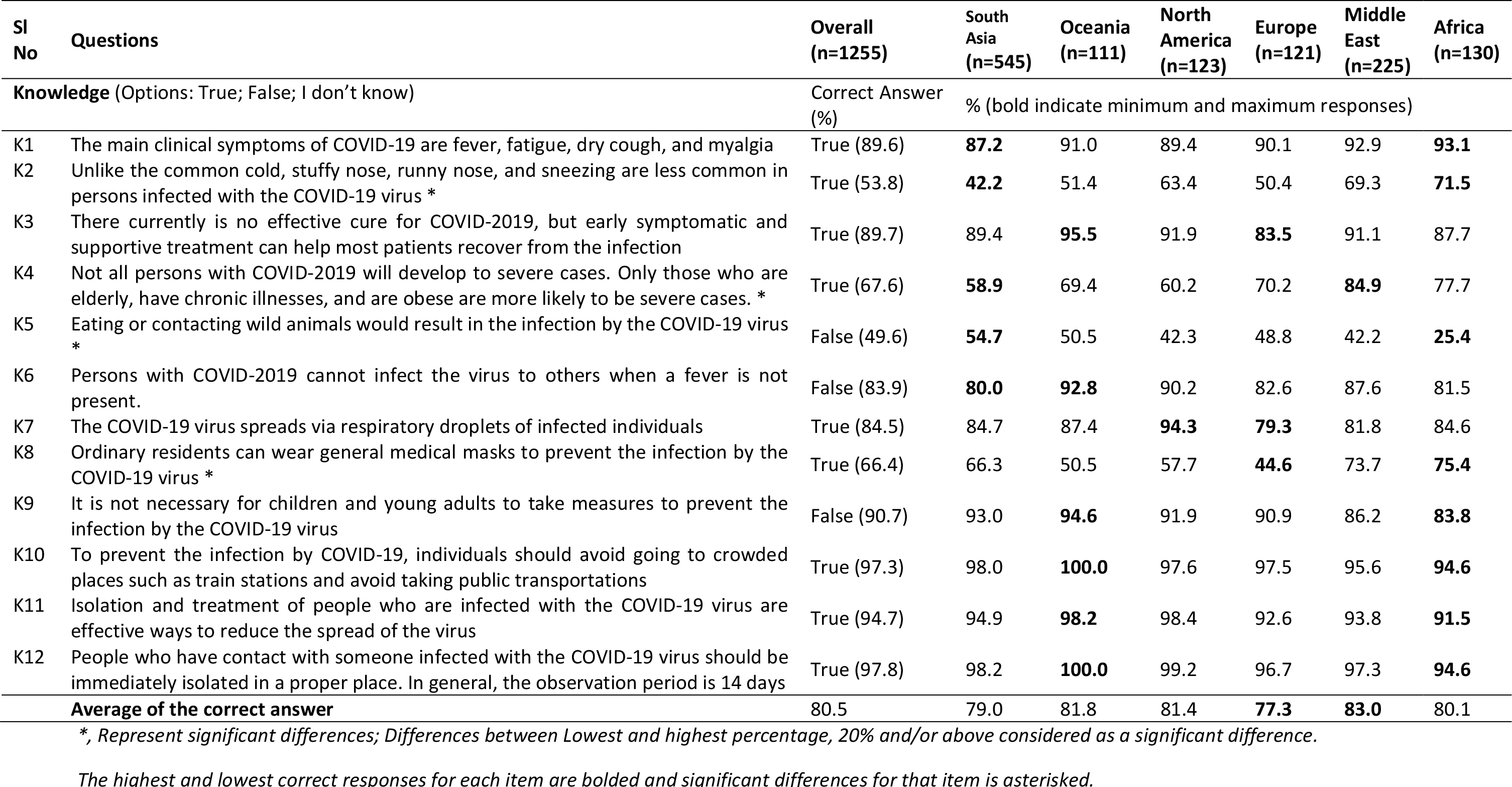
Percentage of correct responses of Knowledge by area of residence.

**Figure 5:**
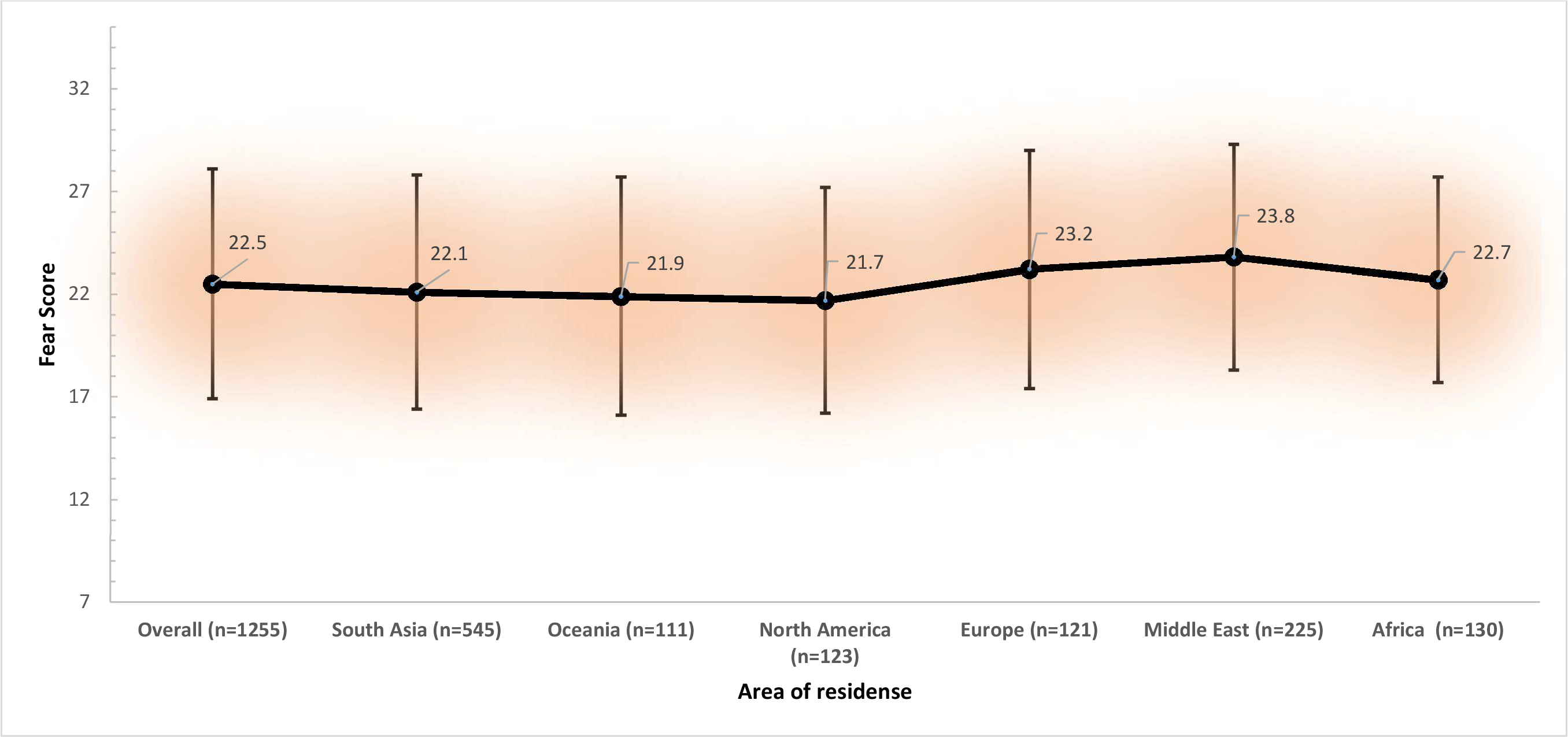
Fear score (mean±SD) distributed by the area of residence.

## References

Adolphs, R., 2013. The Biology of Fear. Curr. Biol. 23, R79–R93. https://doi.org/10.1016/j.cub.2012.11.055

Ahorsu, D.K., Lin, C.-Y., Imani, V., Saffari, M., Griffiths, M.D., Pakpour, A.H., 2020. The Fear of COVID-19 Scale: Development and Initial Validation. Int. J. Ment. Health Addict. https://doi.org/10.1007/s11469-020-00270-8

Ajilore, K., Atakiti, I., Onyenankeya, K., 2017. College students’ knowledge, attitudes and adherence to public service announcements on Ebola in Nigeria: Suggestions for improving future Ebola prevention education programmes. Health Educ. J. 76, 648–660. https://doi.org/10.1177/0017896917710969

Baud, D., Qi, X., Nielsen-Saines, K., Musso, D., Pomar, L., Favre, G., 2020. Real estimates of mortality following COVID-19 infection. Lancet Infect. Dis. https://doi.org/10.1016/S1473-3099(20)30195-X

Bedford, J., Enria, D., Giesecke, J., Heymann, D.L., Ihekweazu, C., Kobinger, G., Lane, H.C., Memish, Z., Oh, M. don Sall, A.A., Schuchat, A., Ungchusak, K., Wieler, L.H., 2020. COVID-19: towards controlling of a pandemic. Lancet. https://doi.org/10.1016/S0140-6736(20)30673-5

Cascella, M., Rajnik, M., Cuomo, A., Dulebohn, S.C., Di Napoli, R., 2020. Features, Evaluation and Treatment Coronavirus (COVID-19), StatPearls. StatPearls Publishing.

Durham, J., 2014. Ethical challenges in cross-cultural research: a student researcher’s perspective. Aust. N. Z. J. Public Health 38, 509–512. https://doi.org/10.1111/1753-6405.12286

Goni, Hasan, Naing, Wan-Arfah, Deris, Arifin, Baaba, 2019. Assessment of Knowledge, Attitude and Practice towards Prevention of Respiratory Tract Infections among Hajj and Umrah Pilgrims from Malaysia in 2018. Int. J. Environ. Res. Public Health 16, 4569. https://doi.org/10.3390/ijerph16224569

Jefferson, T., Foxlee, R., Del Mar, C., Dooley, L., Ferroni, E., Hewak, B., Prabhala, A., Nair, S., Rivetti, A., 2008. Cochrane Review: Interventions for the interruption or reduction of the spread of respiratory viruses. Evidence-Based Child Heal. A Cochrane Rev. J. 3, 951–1013. https://doi.org/10.1002/ebch.291

Jin, J.-M., Bai, P., He, W., Wu, F., Liu, X.-F., Han, D.-M., Liu, S., Yang, J.-K., 2020. Gender Differences in Patients With COVID-19: Focus on Severity and Mortality. Front. Public Heal. 8, 152. https://doi.org/10.3389/fpubh.2020.00152

Johns Hopkins Coronavirus Resource Center, n.d.

Lau, J.T.F., Kim, J.H., Tsui, H., Griffiths, S., 2007. Anticipated and current preventive behaviors in response to an anticipated human-to-human H5N1 epidemic in the Hong Kong Chinese general population. BMC Infect. Dis. 7, 18. https://doi.org/10.1186/1471-2334-7-18

Leung, G.M., Ho, L.-M., Chan, S.K.K., Ho, S.-Y., Bacon-Shone, J., Choy, R.Y.L., Hedley, A.J., Lam, T.-H., Fielding, R., 2005. Longitudinal Assessment of Community Psychobehavioral Responses During and After the 2003 Outbreak of Severe Acute Respiratory Syndrome in Hong Kong. Clin. Infect. Dis. 40, 1713–1720. https://doi.org/10.1086/429923

Markel, H., 1999. Quarantine!: East European Jewish immigrants and the New York City epidemics of 1892.

Person, B., Sy, F., Holton, K., Govert, B., Liang, A., Garza, B., Gould, D., Hickson, M., McDonald, M., Meijer, C., Smith, J., Veto, L., Williams, W., Zauderer, L., 2004. Fear and Stigma: The Epidemic within the SARS Outbreak. Emerg. Infect. Dis. 10, 358–363. https://doi.org/10.3201/eid1002.030750

Rassi, C., Martin, S., Graham, K., de Cola, M.A., Christiansen-Jucht, C., Smith, L.E., Jive, E., Phillips, A.E., Newell, J.N., Massangaie, M., 2019. Knowledge, attitudes and practices with regard to schistosomiasis prevention and control: Two cross-sectional household surveys before and after a Community Dialogue intervention in Nampula province, Mozambique. PLoS Negl. Trop. Dis. 13, e0007138. https://doi.org/10.1371/journal.pntd.0007138

Siddique, M.K. Bin Islam, S.M.S., Banik, P.C., Rawal, L.B., 2017. Diabetes knowledge and utilization of healthcare services among patients with type 2 diabetes mellitus in Dhaka, Bangladesh. BMC Health Serv. Res. 17. https://doi.org/10.1186/s12913-017-2542-3

Squibb, B., Yardley, K., 1999. Playing Healthy, staying healthy: A prevention program for contagious disease. Early Child. Educ. J. https://doi.org/10.1023/A:1022973216955

Survey to Identify the Public Health Ethics Needs of Public Health Practitioners in Canada: Preliminary Results, n.d.

Tachfouti, N., Slama, K., Berraho, M., Nejjari, C., 2012. The impact of knowledge and attitudes on adherence to tuberculosis treatment: A case-control study in a moroccan region. Pan Afr. Med. J. 12. https://doi.org/10.11604/pamj.2012.12.52.1374

Wang, M., Han, X., Fang, H., Xu, C., Lin, X., Xia, S., Yu, W., He, J., Jiang, S., Tao, H., 2018. Impact of Health Education on Knowledge and Behaviors toward Infectious Diseases among Students in Gansu Province, China. Biomed Res. Int. 2018, 1–12. https://doi.org/10.1155/2018/6397340

Yap, J., Lee, V.J., Yau, T.Y., Ng, T.P., Tor, P.-C., 2010. Knowledge, attitudes and practices towards pandemic influenza among cases, close contacts, and healthcare workers in tropical Singapore: a cross-sectional survey. BMC Public Health 10, 442. https://doi.org/10.1186/1471-2458-10-442

Zhong, B.-L., Luo, W., Li, H.-M., Zhang, Q.-Q., Liu, X.-G., Li, W.-T., Li, Y., 2020. Knowledge, attitudes, and practices towards COVID-19 among Chinese residents during the rapid rise period of the COVID-19 outbreak: a quick online cross-sectional survey. Int. J. Biol. Sci. 16, 1745–1752. https://doi.org/10.7150/ijbs.45221

Zhong, B.L., Luo, W., Li, H.M., Zhang, Q.Q., Liu, X.G., Li, W.T., Li, Y., 2020. Knowledge, attitudes, and practices towards COVID-19 among Chinese residents during the rapid rise period of the COVID-19 outbreak: a quick online cross-sectional survey. Int. J. Biol. Sci. 16, 1745–1752. https://doi.org/10.7150/ijbs.45221

